# The French Covid-19 contact tracing app: knowledge, attitudes, beliefs and practices of students in the health domain

**DOI:** 10.1101/2020.10.23.20218214

**Authors:** Ilaria Montagni, Nicolas Roussel, Rodolphe Thiébaut, Christophe Tzourio

## Abstract

**Background:** Many countries around the world have developed mobile phone apps capable of supporting instantaneous contact tracing to control the Covid-19 pandemic. In France, a few people have downloaded and are using the StopCovid contact tracing app. Reasons for this low uptake are unexplored. Students in the health domain are especially concerned and their usage and opinions about the app can inform improvements and diffusion of StopCovid among young people.

**Objective:** To investigate health-related students’ knowledge, attitudes, beliefs and practices about the StopCovid app.

**Methods:** A field survey was conducted among 318 students at the health sciences campus of the University of Bordeaux, France, between September 25^th^ and October 16^th^, 2020. Quota sampling method was used and descriptive statistics were performed.

**Results:** A total of 77.3% (246/318) students had heard about the app, but only 11.3% (36/318) had downloaded it and 4.7% (15/318) were still using it at the time of the survey. Main reasons for not using the app were: belief that it was not effective given its limited diffusion (17.6%, 37/210), lack of interest (17.6%, 37/210) and distrust in data security and fear to be geo-located (15.7%, 33/210). Among those who had not heard about the app, after a brief description of its functioning and confidentiality policy, 52.7% (38/72) would use it. Participants reported that the main solution for increasing the use of the app would be a better communication (71.4%, 227/318).

**Conclusion:** Even among health students, the contact tracing app was poorly used. Findings suggest that improved communication describing its advantages and simplicity of use, and clarifying false beliefs about the app could help improving significantly its uptake.

## INTRODUCTION

While awaiting for effective treatments and vaccines, non-pharmaceutical interventions have been used to contain the spread of the SARS-Cov-2 virus (Kraemer et al., 2020). Besides generalized lockdown, one of the solutions to limit contagion, locate clusters and isolate them is the tracing of infected people. In contact tracing, an index case with confirmed infection is asked to provide information about contact people who were at risk of acquiring infection from the index case within a given time period (between a week and fourteen days) before the positive test result. These contacts are then tracked and advised about their risk, quarantined, and tested (Kretzschmar et al., 2020). Conventional or manual contact tracing is a long process demanding human resources to contact and follow-up people one by one. It can engender several delays and is potentially biased by imperfect recall of contacts (Braithwaite, Callender, Bullock, & Aldridge, 2020). These limitations can be compensated by digital contact tracing (Ferretti et al., 2020; Kretzschmar et al., 2020).

Following the spread of the Covid-19 disease, several apps have been developed across the world to automatically and rapidly trace contacts in real time. Across all continents more than 45 apps are currently used (LI & Guo, 2020). The general functioning of these apps is that each instance running on a mobile device keeps track of the other instances it comes in close contact with. When users inform their app instance they were tested positive, this contact log is used to determine the other instances of the app - and the users, indirectly - that should be notified. Existing apps use different technologies and algorithmic methods to detect contacts between mobile devices (e.g. using short range Bluetooth-Low-Energy information exchange, or GPS-, WiFi-, or Bluetooth-based geo-location), to keep track of these contacts (e.g. using temporary unique identifiers), to evaluate the infection risk (e.g. based on a predicted distance and the duration of the contact), and to notify potentially exposed people using a centralized or decentralized network approach (John Leon Singh, Couch, & Yap, 2020). Efficacy of digital contact tracing systems has been assessed in previous studies thus reinforcing the idea that these tools can help limit the spread of the virus (Ferretti et al., 2020; Kretzschmar et al., 2020; Moreno Lopez et al., 2020; Wilmink et al., 2020).

Nonetheless, national and international statistics show a limited use of these apps (Council of Europe, 2020). Reasons are various: lack of clear communication, fear of confidentiality breach, little testing access, etc. (Dar, Lone, Zahoor, Khan, & Naaz, 2020). In France, the governmental tracing app StopCovid (Government of France, 2020) has been installed more than 2,7 millions of time since the beginning of June (about 6% of the French population, 42 millions), less than the apps of Germany (18 millions), England and Wales (16 millions), and Italy (9 millions). According to national numbers, only 7969 users have declared being Covid-19 positive in the app and only 472 notifications have been sent to potential at-risk contacts as of October 2020. The low uptake of StopCovid explains its lack of effectiveness: the success of app-based contact tracing critically depends on people’s willingness to use the app so as to interrupt the chains of infection transmission. Understanding in depth the levers and barriers for using contact tracing apps can inform on potential improvements as well as public-oriented communication strategies and appropriate political decisions to increase their diffusion.

The recrudescence of the virus after the general lockdown in March-May 2020 has especially concerned young people across France and in the Bordeaux region in particular, where incidence of Covid-19 positive cases among young adults aged 20-30 years has increased to about 252/100,000 per week (week 41-42 Santé publique France, 2020). Since September 1^st^ 2020, several hundreds of students of the University of Bordeaux have been tested and 26 of them returned positive as of November 10^th^. Students in the health domain are on the front line in terms of contagion. First, as many students across France, they are at risk because of their multiple contacts with their peers. They often meet at the University (before, during and after classes), in downtown or for private events. During these encounters, barrier gestures and preventive measures are not always respected. Second, their role as future health-related workers might suppose that they should give the example as users of the app, since they are sensitized to adopt behaviors in favor of health promotion and prevention. Third, most of students in the health domain are in contact with patients directly or indirectly through their interaction with healthcare workers. These different situations, informal, unprotected and in relation with potentially unknown people, are typically those in which contact tracing apps make the most sense. Furthermore, students are digitally literate and are supposed to be more at ease with download and use of apps. French students in the health domain are thus a priority target for the uptake of the StopCovid app.

The aim of this study was to describe knowledge, attitudes, beliefs and practices (KABP) about the StopCovid app among students in the Bordeaux region, France. Reasons for downloading or not the app and fake news concerning its functioning were also explored.

## METHODS

### The StopCovid app

StopCovid was launched by the French Government the 2^nd^ June 2020. It was developed by a team of public and private partners lead by the French National Institute for Research in Digital Science and Technology (Inria), and is available in both Apple and Google Play stores as it works on iOS and Android phones. The app is based on Bluetooth signals, running in the background of the phone with low-energy wireless transmission (Cunche, Boutet, Castelluccia, & Lauradoux, 2020). Once the app is activated, the phone logs other phones it comes in contact with, assuming concerned devices are running StopCovid. These logs do not include any identifying information about the user: they used random ID codes that change every 15 minutes and get trashed completely once they are older than 14 days (the incubation period for Covid-19). The app does not locate the user (no GPS-, WiFi- or Bluetooth-based geolocation); it only knows which random IDs the phone has come into contact with. Transparent and anonymous, the app does not collect any personal data nor contact details. If a user declares being positive for Covid-19 using a code delivered with the test results, the app will send that record of the rotating IDs to a server, which in turn will send them out to other devices using the system (Gorce, Egan, & Gribonval, 2020). Anyone who has been nearby in the last two weeks will be pinged with an alert. More precisely, this notification is sent if the person has spent more than 15 minutes within 1 meter of an index case. Users are then recommended to inform their general practitioner, get tested and self-isolate, thus potentially stopping another line of transmission. A new version of the app has been available since the 22nd Octobre, 2020. The main changes are i) the name of the app “TousAntiCovid” ii) the availability of news in regards of the COVID-19 that are regularly updated.

### The field survey

This study was conducted within the framework of the larger ongoing i-Share cohort study (Internet-Based Students Health Research Enterprise, www.i-share.fr), a French nationwide Web-based survey on the health and well-being of university students, whose principal investigators and operational staff are based at the University of Bordeaux (Montagni, Qchiqach, Pereira, Tully, & Tzourio, 2018).

The field survey consisted in a paper questionnaire administered face-to-face by five undergraduate students (interviewers) who had been trained to take notes, fill in the questionnaire and describe the app to respondents. Interviewers approached their peers in the halls, canteen, courtyards, library and study rooms at the health sciences campus of the University of Bordeaux. The collection of the data started on September 25^th^ 2020 and ended on October 16^th^ 2020. A sample size of 300 respondents was targeted with quotas set for the sample to be representative of the overall population of students in the health domain at the University of Bordeaux (n=16,566) in terms of sex, age (18-30), specific field of health-related study (medicine, dentistry, nursing, pharmacy, public health, etc.) and year of study (1 to >6 years) (see Table 1 for the details). Inclusion criteria were: being aged >18 years, being a student in the health domain enrolled at the University of Bordeaux, and providing oral informed consent.

**Table 1.** Sociodemographic characteristics of the study population (n=318) and comparison with all students in the health domain (n=18 000)

|  |  | Study population<br>(n=318) |  | Total health-related student<br>population at the<br>University of<br>Bordeaux<br>(n=16,566) |  |
| --- | --- | --- | --- | --- | --- |
|  |  | n | % | n | % |
| **Sex** |  |  |  |  |  |
|  | Female | 209 | 65.7 | 11713 | 70.7 |
|  | Male | 109 | 34.3 | 4853 | 29.3 |
| **Age** |  |  |  |  |  |
|  | Mean (SD) | 20.4 | (2.39) | 23.8 | NA |
| **Year of study** |  |  |  |  |  |
|  | 1 | 129 | 40.6 | 4445 | 31.2 |
|  | 2 | 64 | 20.1 | 2341 | 16.4 |
|  | 3 | 46 | 14.5 | 2334 | 16.4 |
|  | 4 | 23 | 7.2 | 1200 | 8.4 |
|  | 5 | 44 | 13.8 | 1267 | 8.9 |
|  | >5 | 10 | 3.1 | 2651 | 18.6 |
|  | Other | 2 | 0.6 | NA | NA |
| **Field of study** |  |  |  |  |  |
|  | Medicine | 193 | 60.7 | 7104 | 49.9 |
|  | Pharmacy | 61 | 19.2 | 1138 | 8.0 |
|  | Dentistry | 12 | 3.8 | 521 | 3.7 |
|  | Nursing | 14 | 4.4 | 3972 | 27.9 |
|  | Public Health | 15 | 4.7 | NA | NA |
|  | Other | 23 | 7.2 | 1503 | 10.6 |

The questionnaire was co-designed and tested by a team of 14 public health researchers and operational staff following a structured survey construction method in 5 steps (Pazzaglia, Stafford, & Rodriguez, 2016). The final questionnaire was composed of 36 items, 14 of which were common to all students (socio-demographic characteristics, suggestions for increasing the diffusion of the app, willing to recommend the app to family and friends, and fake news about data collection and sharing within the app). The other items were administered based on four different scenarios: (1) the student has already heard about the app and has downloaded it; (2) the student has already heard about the app and has not downloaded it; (3) the student has never heard about the app but would download it; and (4) the student has never heard about the app and would not download it. Specific questions were then asked depending on the scenario. Before answering further questions, students who had not heard about the app were provided a brief description of it. After having responded to fake news about data collection and sharing within the app, all students were given the correct answers. The English version of the questionnaire is available as *Supplemental File*. The time of administration and completion of the questionnaire was about 10 minutes. The field survey was approved by the University of Bordeaux. The oral informed consent reassured students of the anonymous format of the survey and use of collected data for research purposes only.

All data from the paper questionnaires were entered by the student interviewers in a digital database through the EpiData® software version 3.1. A descriptive analysis was performed, presenting all variables and measures in the form of numbers and percentages for qualitative variables and mean and standard deviation (SD) for quantitative variables. Chi2 or exact Fisher frequency comparison tests were used to identify statistically significant differences by age, gender, field and year of study, modified, if necessary, to binary variables. Statistical significance was defined with a p-value < 0.05. The data were analyzed with SAS® version 9.3(SAS Institute, Cary, NC, USA).

## RESULTS

A total of 590 students were approached to complete the survey after a brief explanation of its objective: 318 completed the survey while 272 refused to participate for a final participation rate of 53.9%. Reasons for not participating were lack of time or no interest for the topic of the study. The final study sample was composed of 65.7% (209/318) female students and was aged 20.4 years (2.39 SD). All fields and years of study were represented. Medical students were the most numerous, 60.7% (193/318), in line with the total number of medical students at the University of Bordeaux. In accordance with University-related statistics, first year students were also the most represented (40.6%, 129/318). Figure 1 shows the flowchart of the study population and Table 1 the sociodemographic characteristics with the corresponding data for the total population of health-related students at the University of Bordeaux.

**Figure 1.**
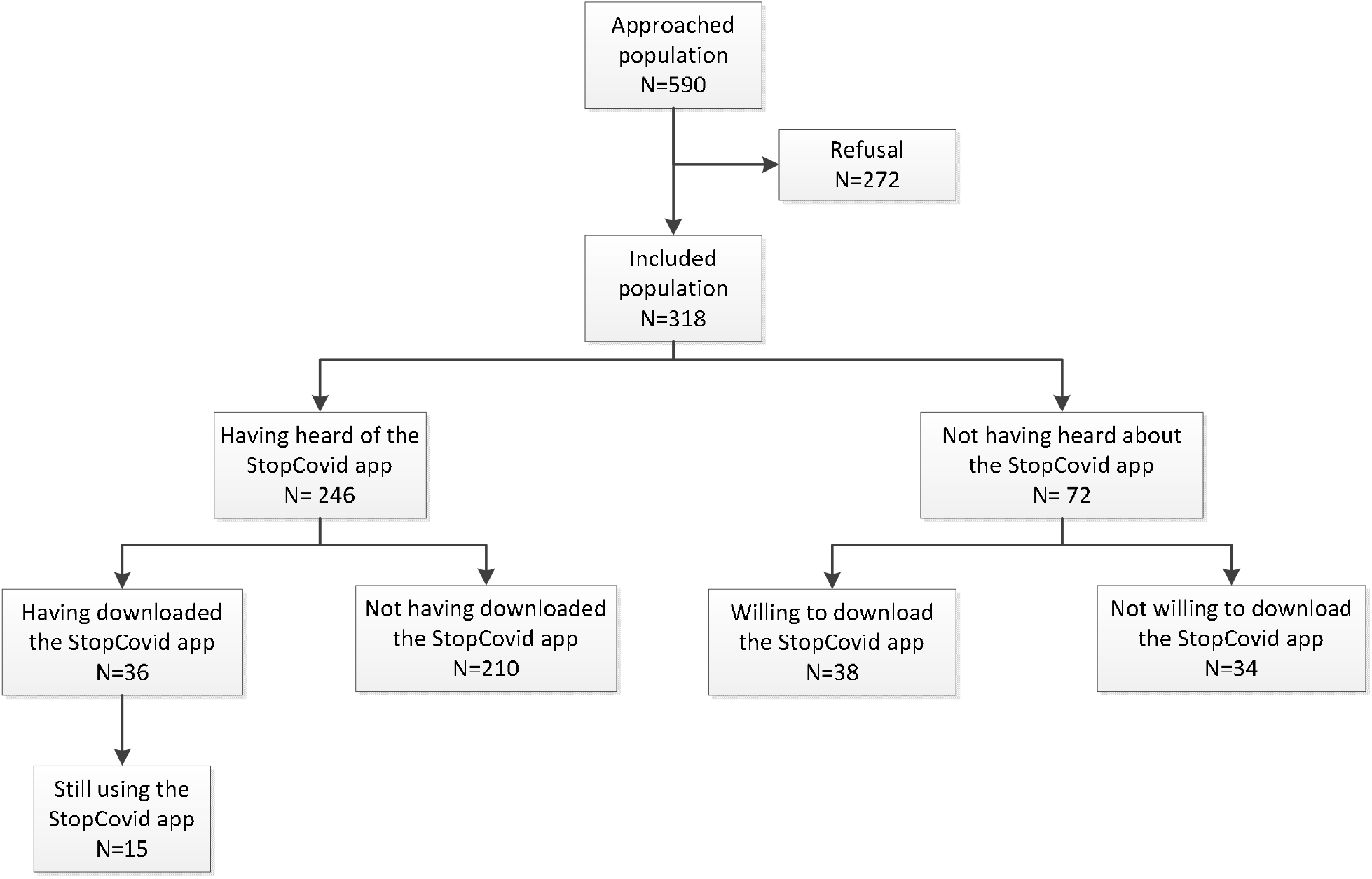
Flowchart of the study population (n=318)

The majority of participants had already heard about the app (77.3%, 246/318), mostly through the media (87.8%, 216/246) and secondly through family and friends (15.9%, 39/246). Concerning these variables, no statistically significant differences were found based on age, sex, field and year of study.

Most of the students correctly knew that the app was promoted by the government (72.8%, 179/246), but 25.2% (62/246) answered that they did not know who the promoter was. Male students knew significantly more than female students that the app was promoted by the government (81.2% vs. 68.9%; p-value = 0.0313). Female students were significantly more likely to ignore the promoter of the app compared to male students (29.2% vs. 17.6%; p-value = 0.0473). Students of any health-related discipline than medicine responded significantly more than medical students that the application was promoted by a research laboratory (3.9% vs. 0.0%; p-value = 0.0297). Medical students were significantly more likely to ignore the promoter of the app (30.1% vs. 18.4%; p-value = 0.0383). No statistically significant differences were found based on age and year of study.

Among those who had heard about the app, 14.6% (36/246) had actually downloaded it when it was first released in June (61.1%, 22/36) or with the new cases of Covid-19 at the beginning of the University year (16.7%, 6/36). Of them, 41.6% (15/36) were still using the app. Of the total sample, students using the app at the moment of the survey were 4.7% (15/318), while those having uninstalled the app had used it from one day (16.7%, 6/36) to several weeks (16.7%, 6/36). Main reasons for uninstalling the app were that it was not so useful (66.7%, 14/21), the respondent forgot to activate the Bluetooth (23.8%, 5/21), the app drained the phone battery (19.0%, 4/21) and too few people were using it thus making the app ineffective (19.0%, 4/21). Accordingly, students reported that the main default of the app was that it seemed inefficient given its limited uptake (48.6%, 17/35, 1 missing). For 25.7% (9/35) the app presented technical problems like draining the battery, depending on Bluetooth or occupying too much space on the phone. Concerning all previous variables, no statistically significant differences were found based on age, sex, field and year of study.

These students also reported that its qualities were that it was easy to use (51.4%, 18/35) and that it was reassuring (25.7%, 9/35). Male students found the application significantly more user-friendly than female students (70.6% vs. 33.3%; p-value = 0.0275). Female students recognized several qualities of the application significantly more than male students (38.9% vs. 5.9%; p-value = 0.0408; statistical power = 0.527). Concerning this variable on the quality of the app, no statistically significant differences were found based on age, field and year of study.

Reasons for downloading or not the app are showed in Table 2 (multiple answers possible for each individual).

**Table 2.** Reasons for downloading or not the StopCovid app.

|  |  | Yes, I have downloaded the<br>app (n=36) |  | Yes, I would download<br>the app (n=38) |  |
| --- | --- | --- | --- | --- | --- |
|  |  | n | % | n | % |
| **Reasons for downloading the app** |  |  |  |  |  |
|  | Out of curiosity | 13 | 36.1 | 14 | 36.8 |
|  | To protect my family, others and myself from possible infection | 13 | 36.1 | 24 | 63.2 |
|  | It was the government that advised the downloading of the app | 5 | 13.9 | 0 | 0.0 |
|  | The app could be useful to contain the spread of the virus in general | 5 | 13.9 | 200 | 52.6 |
|  | Am afraid of the virus and all strategies are good to avoid it | 1 | 2.8 | 1 | 2.6 |
|  | Am reassured the app is anonymous | NA | NA | 0 | 0.0 |
|  | Other | 11 | 30.6 | 1 | 2.6 |
|  |  | No, I have not downloaded the<br>app (n=210) |  | No, I would not download<br>the app (n=34) |  |
|  |  | n | % | n | % |
| **Reasons for not downloading the app** |  |  |  |  |  |
|  | Cannot see the interest or need | 90 | 42.9 | 17 | 50.0 |
|  | Do not like the general idea of this app | 10 | 4.8 | 1 | 2.9 |
|  | Do not know how it works, did not get enough information on the app | 23 | 11.0 | 2 | 5.9 |
|  | Am suspicious of this type of app | 18 | 8.6 | 1 | 2.9 |
|  | Do not trust, because do not know who is offering this app:<br>industry etc. | 11 | 5.2 | 1 | 2.9 |
|  | Not sure about the security of the data, fear of being geo-located | 33 | 15.7 | 1 | 2.9 |
|  | My family and friends have discouraged me from downloading it | 1 | 0.5 | 0 | 0.0 |
|  | No space on my phone or it is not powerful enough to have an<br>extra app (battery, Bluetooth) | 27 | 12.9 | 6 | 17.6 |
|  | Do not carry my phone with me at all times | 2 | 1.0 | 4 | 11.8 |
|  | Do not use public transportation and/or do not go out much in<br>public places (do not come into contact with strangers) | 11 | 5.2 | 4 | 11.8 |
|  | It does not seem to be effective (too few people use it) | 37 | 17.6 | 8 | 23.5 |
|  | Heard negative feedback on this app | 7 | 3.3 | 1 | 2.9 |
|  | Do not really have time to think about it | 37 | 17.6 | 3 | 8.8 |
|  | By negligence, not concerned | 28 | 13.3 | 5 | 14.7 |
|  | Not sure what it is all about, the principle and or the functioning | 11 | 5.2 | NA | NA |
|  | Other | 15 | 7.1 | 2 | 5.9 |

Among those having heard about the app, but who had not downloaded it, main reasons for not using the app were: lack of interest (42.9%, 90/210), belief that it was neither effective nor useful given its limited diffusion (17.6%, 37/210), and distrust in data security and fear to be geo-located (15.7%, 33/210). The majority of students having heard about the app but who had not downloaded it might change their mind and use the app if they had more information about it through better communication strategies (29.0%, 61/210) and if more people would use it (25.7%, 54/210). Nonetheless, 26.2% (55/210) would not change their mind and still not download the app. On the other hand, main reasons for downloading the app were out of curiosity (36.1%, 13/36) and to protect one’s family, others and oneself from possible infection (36.1%, 13/36).

Students who had never heard about the app (n=72) were asked to imagine its content and objective: 41.7% (30/72) reported that it was an app providing advice and information about Covid-19; 29.2% (21/72) that it was an app to limit the spread of the virus; 29.2% (21/72) did not know; and 15.3% (11/72) answered “other”. After a short description of the app, 52.7% (38/72) said they would download it. Reasons for downloading or not the app are reported in Table 1 and are similar to those provided by the sample having heard about the app. Among these students who had never heard about the app and who were still not willing to download it after a brief description (n=34), 32.4% (11/34) would not change their mind, 17.6% (6/34) would download it if more people used it, and 11.8% (4/34) would download it if they had a more performing telephone.

Concerning the functioning of the app, 83.3% (30/36) of the respondents said that they were able to explain it. However, when further asked about geo-location, access to contact information and how data were transmitted and stocked, their answers were mostly incorrect. As expected, students not having heard about the app before, but who were presented a quick description of it during the survey, provided correct answers more than their peers. Detailed results are shown in Table 3.

**Table 3.**
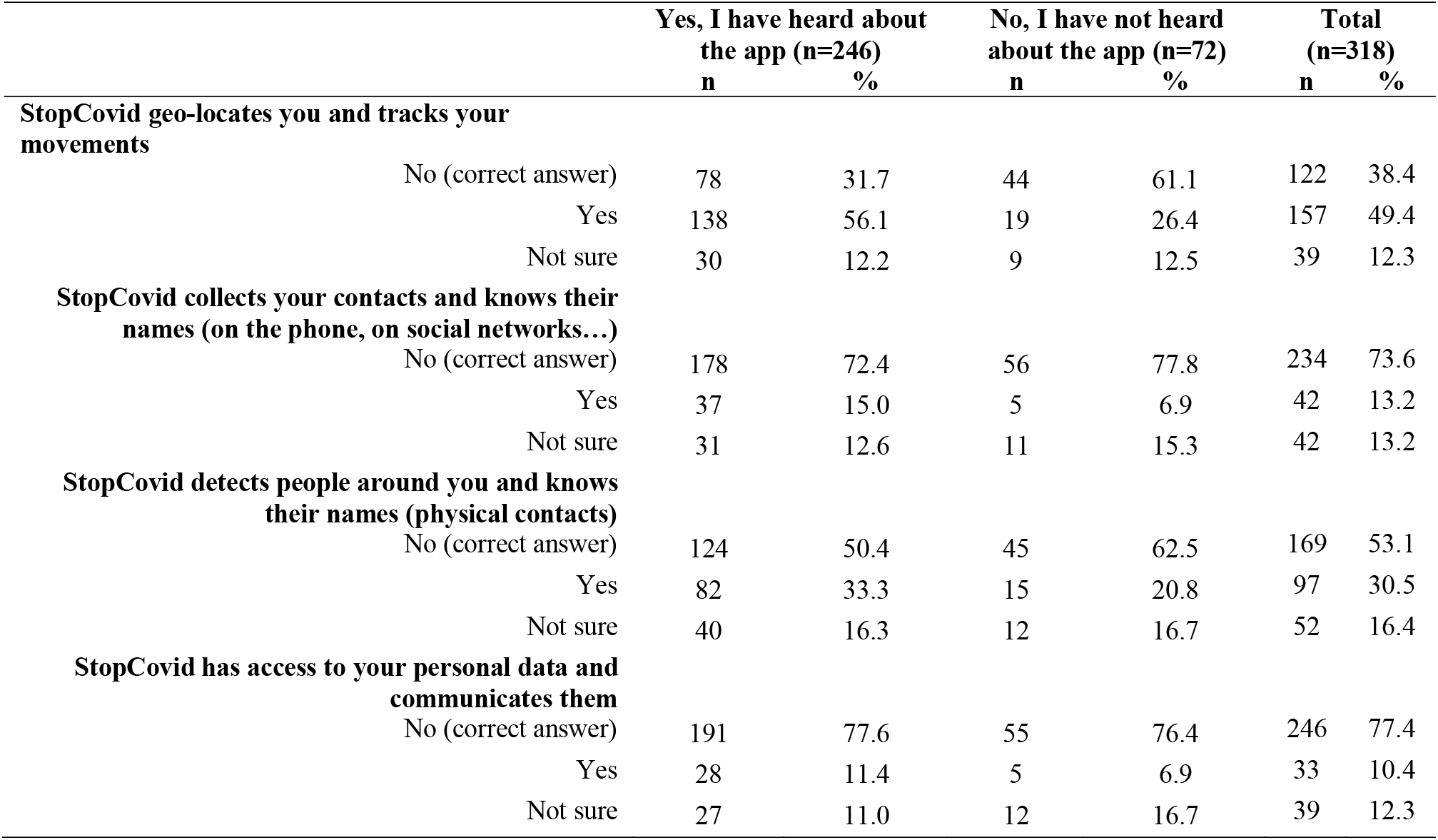
Knowledge about the functioning and data management of the StopCovid app.

|  |  | Yes, I have heard about<br>the app (n=246) |  | No, I have not heard<br>about the app (n=72) |  | Total<br>(n=318) |  |
| --- | --- | --- | --- | --- | --- | --- | --- |
|  |  | n | % | n | % | n | % |
| **StopCovid geo-locates you and tracks your movements** |  |  |  |  |  |  |  |
|  | No (correct answer) | 78 | 31.7 | 44 | 61.1 | 122 | 38.4 |
|  | Yes | 138 | 56.1 | 19 | 26.4 | 157 | 49.4 |
|  | Not sure | 30 | 12.2 | 9 | 12.5 | 39 | 12.3 |
| **StopCovid collects your contacts and knows their names (on the phone, on social networks...)** |  |  |  |  |  |  |  |
|  | No (correct answer) | 178 | 72.4 | 56 | 77.8 | 234 | 73.6 |
|  | Yes | 37 | 15.0 | 5 | 6.9 | 42 | 13.2 |
|  | Not sure | 31 | 12.6 | 11 | 15.3 | 42 | 13.2 |
| **StopCovid detects people around you and knows their names (physical contacts)** |  |  |  |  |  |  |  |
|  | No (correct answer) | 124 | 50.4 | 45 | 62.5 | 169 | 53.1 |
|  | Yes | 82 | 33.3 | 15 | 20.8 | 97 | 30.5 |
|  | Not sure | 40 | 16.3 | 12 | 16.7 | 52 | 16.4 |
| **StopCovid has access to your personal data and communicates them** |  |  |  |  |  |  |  |
|  | No (correct answer) | 191 | 77.6 | 55 | 76.4 | 246 | 77.4 |
|  | Yes | 28 | 11.4 | 5 | 6.9 | 33 | 10.4 |
|  | Not sure | 27 | 11.0 | 12 | 16.7 | 39 | 12.3 |

Finally, the whole sample (n=318) was asked about the factors increasing the use of the app. For the majority of them (71.4%, 227/318) the solution was a better communication strategy. Other factors were: making the app compulsory (14.2%, 45/318); registering more Covid-19 cases (9.4%, 30/318); more information and explanations about the app (6.6%, 21/318); better technical features (3.1%, 10/318); and “other” (20.1%, 64/318).

## DISCUSSION

### Principal findings and interpretation

One student out of five had never heard about the StopCovid app: this rate is surprisingly high considering that students in the health domain should be informed of existing tools to limit the spread of coronavirus. Of the students having heard about the app, mostly through the media, 15% had actually downloaded the app and, in the whole sample, only about 5% were still using it at the time of the survey. These percentages confirm the low uptake of the StopCovid app as reported by national statistics. Main reasons for not downloading the app were that it was deemed poorly interesting and ineffective since very few people were using it. False beliefs were also present: one student out of six had not downloaded the app for fear of being geo-located and that personal data could be collected and shared. Indeed, half of the participants, mostly including those having already heard about the app, falsely believed that the app was intrusive and had access to phone contacts. Among those who received a clear explanation of the functioning of the app and its confidentiality policy (no geo-location, no access to personal data), one student out of two felt reassured and would finally download the app. This result suggests that simple, clear and straightforward information might increase app-related knowledge and acceptability. Accordingly, when asked how they would improve StopCovid use, 71% of students suggested deploying better communication strategies to increase the uptake of the app especially rising its interest.

Our study shows that, while officials in charge at different levels of promoting the app and experts gloss over the reasons for non-use of this app by the population, one of the main reasons of this low uptake is the lack of a simple and factual communication delivered appropriately. Even in health students who are expected to be highly aware of the tools to prevent this epidemic and to be exposed to quality information, the information they had about this app was poor and fraught with false beliefs. On the positive side, clear and straightforward information should increase app-related knowledge and acceptability.

### Comparison with prior work

A multicountry study, including 996 adults from France, was conducted to explore acceptability of and intention to use a contact-tracing app concerning Covid-19 (Altmann et al., 2020). Differently from our survey, questions about future behavior related to the app were hypothetical and not based on a concrete existing contact tracing digital tool: participants could not visualize how such a system works and had not had any experience of it. This study found strong support for the general idea of this type of app: almost 75% of respondents said they would probably or definitely download the app. Thus, these results are more optimistic than the findings of our study where the real uptake of the app was very limited. However, when provided with clear explanation of the app, one student out of two was convinced to download the app within our study. Differences between the results of our study and those of the multicountry study might be explained by the study population (general population vs young people) and the fact that the app presented in the multicountry study was hypothetical vs a real app. Furthermore, in the multicountry study, concerns about cybersecurity and privacy, together with a lack of trust in the government, were mentioned as the main barriers to adoption. Distrust in data security and fear of being geo-located were also reported by our sample, even if not as the principle reason not to use the app. The main reason for not using the app was its potential lack of effectiveness. Controlling Covid-19 requires a high population uptake of automated contact-tracing apps (estimates from 56% to 95%) (Braithwaite et al., 2020). Participants to this study underlined the need for sufficient population uptake to make the app useful and successful. Up to the present, the diffusion of StopCovid is limited thus representing an obstacle to its actual efficacy.

Another study on motivations for adopting a contact tracing app use was conducted with 406 German-speaking participants (Kaspar, 2020). Results showed that younger respondents were more willing to use the app, which strengthens the idea that students are a suitable target for this type of digital interventions. Young people are usually very likely to carry their phone with them, which might explain their easiness to download and use apps in general. Frequent usage of mobile phones might be related to a stronger perception of the potential benefits of the app. Similarly to the previous multicountry study, the perceived severity of and vulnerability to data misuse were negatively related to the participants’ motivation for using a contact tracing app. The participants’ general trust in official app providers was the most important independent variable with respect to app use motivation. This result confirms once again the significant role of data security issues and trust in the context of app-based measures to combat the current pandemic. Internationally, the issue of confidentiality of data transmitted by contact tracing apps has been raised as the most important barrier to download and use the app (Parker, Fraser, Abeler-Dörner, & Bonsall, 2020).

A study conducted among 1500 Belgians (Walrave, Waeterloos, & Ponnet, 2020) on use of a hypothetical contact tracing app also reported that the clarity of the functioning of the app was correlated to the will to use it. This result is in line with our findings. Uptake of the app is strictly related to the understanding of its contents and objectives. When students understand that the app does not follow them nor access their personal information, they are more willing to download it. In fact, in our sample, intention to use the app increased with the access to more and clearer information: participants wished the app to be better promoted and explained.

A sample of 900 Dutch adults (Jonker et al., 2020) was questioned on their will to adopt a hypothetical contact tracing app and the predicted adoption was of 64.1%. This rate strongly varied by age group: the adoption rates of the app ranged from 45.6% to 79.4% for people in the oldest and youngest age groups (i.e.,≥75 years vs 15-34 years), respectively. Educational attainment, the presence of serious underlying health conditions, and the respondents’ stance on Covid-19 infection risks were also correlated with the predicted adoption rate. Once again, younger adults proved to be more prone to download this type of app.

Finally, concerning experience with contact tracing apps in Asia, Singapore attracted considerable early notice for its pioneering role in developing a Covid-19 contact tracing app based on Bluetooth, TraceTogether, with a 20% adoption rate and public discussion of the privacy implications determining users’ lack of motivation to download the app (Goggin, 2020). Furthermore, the app received criticism for it drained the phone battery, which is one of the defaults also reported in this study about the StopCovid app. Privacy issues are the main issue raised for the South Korean contact tracing app, confirming that cybersecurity and privacy are to be considered as a strong barrier for adopting this type of app internationally (Park, Choi, & Ko, 2020).

### Strengths and Limitations

This was one of the first studies reporting data on students’ KABP about a contact tracing app in a pandemic context. Previous studies have explored the intention of downloading this type of apps as a general idea, but were not based on a developed and currently diffused app (Altmann et al., 2020; Jonker et al., 2020; Kaspar, 2020; Walrave et al., 2020). Reasons for downloading and using the app were explored in depth to inform future steps to increase its diffusion. The specific focus on students was another strong point of this study: young people are especially concerned by the transmission of the virus in subsequent coronavirus waves. Mobilizing this population to adopt the app is key in this particular epidemiological context.

Limitations of the study include the relatively small sample. More than 300 students in the health domain were interviewed among a total population of 18,000 students. Findings cannot be generalized, but the sample was recruited according to quota sampling in order to be, as much as possible, representative of sex, age, specific field of study (from medicine to pharmacy) and year of study. This might increase the representativeness of the interviewed population group. The small sample also justifies the few significant differences that were identified. This is confirmed by the low statistical powers that were obtained following performed statistical tests (< 0.50).

### The new version of the StopCovid app

StopCovid has recently received several criticisms and even the French Prime Minister Jean Castex has officially declared not having downloaded the app. The government has considered the low uptake of the app as the main issue of StopCovid. Some weeks after the implementation of this study, the French president Emmanuel Macron has announced the launch of a new contact tracing app, TousAntiCovid with new features: provision of information on new cases, effective R, incidence rate, positivity rate, occupancy rate; app registrations, reported cases, app alerts; and testing locations. Thus, this study concerned the first version of this app with relatively small changes and same contact trading functioning compared to the new version. Obtained results might be easily transposed to the new TousAntiCovid app.

### Implications

This survey was conducted as the preliminary phase of a complex intervention aimed at promoting the uptake of the StopCovid app among students in the health domain at the University of Bordeaux. After this first appraisal of KABP about StopCovid, the next steps are to implement a series of actions on the premises of the University. Professors and lecturers have been mobilized and trained to present StopCovid to their students during classes. Furthermore, students will be informed also by other communication supports like short videos on the University website and intranet, flyers, posts on social networks, posters, etc. Student ambassadors and associations will also be involved in the diffusion of the app. This complex intervention will be evaluated through a second series of random field surveys aimed at observing an increase in the number of downloads of the app. Depending on the results of the evaluation, the intervention will be extended to students of other fields of study at the University of Bordeaux and in other Universities across France.

## Conclusion

Overall, broad support for app-based contact tracing was found notwithstanding its low uptake. Results suggest that the functioning and purpose of the StopCovid app were not well known and appraised among students, especially because of the clear lack of factual communication about it. Efforts are to be taken in these terms in order to increase knowledge about the app and diffuse its adoption in the young who represent a priority target audience. The way the app traces contacts should be better explained so as to maximize download and consequential usage of the app by eliminating any potential false belief.

## Supporting information

Questionnaire

## Data Availability

All data generated or analysed during this study are included in this manuscript. The full dataset is available on reasonable request.

## ACKNOWLEDGEMENTS

The project is led by the University of Bordeaux, in close collaboration with the University Hospital (CHU) of Bordeaux, the Regional Health Authority (ARS), and the National Institute for Research in Digital Science and Technology (Inria).

The authors wish to thank the i-Share team (www.i-share.fr) for managing and conducting the study and for revising the questionnaire: Julie Arsandaux, Fadi El Khoury, Shérazade Kinouani, Mélissa Macalli, Elena Milesi, Marie Mougin, Garance Perret and Clothilde Pollet. Within the i-Share team, the authors wish to thank especially Edwige Pereira and Aude Pouymayou for their support for statistical analysis. The authors are also indebted to the student ambassadors that carried out the field survey: Idriss Boukili, Justine Char, Jade Giacosa, Oleg Hounkpatin, Jeanne Langlois and Marie Lanneluc.

## Author approval

All authors have seen and approved the manuscript

## Authors’ Contributors

IM was responsible for writing, literature search, data collection and interpretation. CT, RT and NR were responsible for study design and revision of the manuscript.

