## Supplementary material for "The French Covid-19 contact tracing app: knowledge, attitudes, beliefs and practices of students in the health domain": Questionnaire

**SUPPLEMENTAL FILE**

| **English version of the StopCovid field survey questionnaire** |
| --- |

**ID number:**

**Date:**

**Place:**

**Name of the interviewer:**

**1) Do you agree to have your answers registered and coded to be analyzed in a totally anonymous way by a team of researchers from the University of Bordeaux?** 1=YES 0=NO

If the student answers "NO", the survey ends. Note the refusal.

Reason for refusal: ________________________

**2) Are you a student in a health program at the University of Bordeaux?** 1=YES 0=NO

**3) Field of study (medicine, pharmacy, dentistry, speech therapy, etc.):**
**__________________________**

**4) Year of study:** 1=1 2=2 3=3 4=4 5=5 6=>5 7=Other

If "Other" specify: ______________________________________

**5) Age (between 18 and 30): __ __**

**6) Gender:** 1=Male 2=Female

**7) What is the operating system of your phone?** 1=iOS-Apple 2=Android

**8) Have you already heard about the StopCovid mobile app?** 1=YES 0=NO

**SCENARIO 1 "Yes, I have already heard about the StopCovid mobile app**

**9) How did you hear about it?**

1. Media (press, social networks, television...)
2. Family and friends
3. At the University (teachers, Student Health Centre, student associations...)
4. Other

If "Other", specify: ____________________________

**10) Do you know who promotes this app?**

1. A private company
2. A research laboratory
3. The Government
4. Hospital or other health care institution
5. Do not know
6. Other

If "Other", specify: ___________________________

**11) Have you downloaded the app?** 1=YES 0=NO

| IF "YES, I HAVE DOWNLOADED THE APP" | IF "NO, I HAVE NOT DOWNLOADED THE APP" |
|---|---|
| **11.1) When did you download it?**<br>1. As soon as it was released, early June<br>2. With the new cases of COVID-19 this summer<br>3. At the beginning of the University year<br>4. Cannot remember<br>5. Other<br>If "Other", specify: ____________________<br><br>**11.2) Why did you download it?**<br>1. Out of curiosity<br>2. To protect my family, others and myself from possible infection<br>3. It was the government that advised the downloading of the app<br>4. The app could be useful to contain the spread of the virus in general<br>5. Am afraid of the virus and all strategies are good to avoid it<br>6. Other<br>If "Other", specify: ____________________<br><br>**11.3) Can you explain how the app works?**<br>1=YES 0=NO 2=DO NOT KNOW<br>*#Ask the student to explain how the app works. Check that the explanation is close to this description: The app must be installed. Then it must be launched. Once launched, it can be disabled/enabled (a button with disable/enable is used in StopCovid to suspend/restart contact* | **11.7) Why did not you download it?**<br>1. Cannot see the interest or need<br>2. Do not like the general idea of this app<br>3. Do not know how it works, did not get enough information on the app<br>4. Am suspicious of this type of app<br>5. Do not trust, because do not know who is offering this app: industry etc.<br>6. Not sure about the security of the data, fear of being geo-located<br>7. My family and friends have discouraged me from downloading it<br>8. No space on my phone or it is not powerful enough to have an extra app (battery, Bluetooth)<br>9. Do not carry my phone with me at all times<br>10. Do not use public transportation and/or do not go out much in public places (do not come into contact with strangers)<br>11. It does not seem to be effective (too few people use it)<br>12. Heard negative feedback on this app<br>13. Do not really have time to think about it<br>14. By negligence, not concerned<br>15. Not sure what it is all about, the principle and or the functioning<br>16. Other<br>If "Other", specify:_____________________ |

*tracing). Bluetooth must be activated. It can also be stopped and uninstalled.#*

**11.4) How long did you use it?**

1. One day, just for testing, then did not use it anymore
2. Between one day and a week, then did not use it anymore
3. For more than a week, then did not use it anymore
4. Still use it (even if uninstalled and reinstalled)
5. Cannot remember

**11.4.1) Why don't you use it anymore?**

1. Took up too much space on my phone
2. Using Bluetooth all the time was annoying
3. Consumed a lot of battery power
4. Not so useful, no longer saw any interest in having it
5. Did not want my contacts to be traced
6. Did not have a phone all the time, so it cannot be efficient
7. Did not work well
8. Did not think to activate the Bluetooth
9. Did not understand how it works
10. Too few people use it, so it cannot be effective
11. Other

If "Other", specify: ____________________

**11.5) The main quality of the app?**

1. It is easy to use
2. It seems to be effective
3. It is reassuring
4. It does not use personal data or geo-location
5. Other

If "Other", specify : ____________________

**11.6) The main default of the app?**

1. It is not easy to use
2. It does not seem to be effective (few people use it)
3. It poses technical problems (battery, Bluetooth, place on the phone...)
4. Not totally sure of its transparency and anonymity
5. Other

*#If the student says he or she is not familiar with the app, explain it to him or her quickly#*

**11.8) What could make you change your mind and download the app?**

1. Have a better phone
2. Learn more about the app (with better communication, for example)
3. Have positive feedback on its usefulness
4. Having cases of COVID-19 among my family and friends
5. Be even more reassured about its transparency and the protection of anonymity (clearer information on the transmission of information)
6. Many more people who use it
7. Know that the app is used by a majority of the people around me (e.g. other students that I may come across)
8. Nothing would change my mind
9. Other

If "Other, specify:______________________

| |
|---|
| If "Other", specify : ____________________ |

For both groups of SCENARIO 1

**12) In your opinion, what could help diffuse the app more widely?**

1. A better communication strategy (more visibility, clearer messages)
2. Providing better technical features (battery, Bluetooth, space on the phone)
3. An explosion of Covid-19 cases
4. Making it mandatory
5. Do not know
6. Other

If "Other", specify: _________________________

**13) Would you advise those around you to download the app?** 1=YES 0=NO 2=DO NOT KNOW

**14) Which of the following statements are true or false? You can also answer that you are not sure of the answer.**

**14.1) StopCovid geo-locates you and tracks your movements:** 1=True 0=False 2=Not sure

**14.2) StopCovid collects your contacts and knows their names (telephone directory, contacts on social networks...):** 1=True 0=False 2=Not sure

**14.3) StopCovid detects people around you and knows their names (physical contacts):** 1=True 0=False 2=Not sure

**14.4) StopCovid has access to your personal data and communicates them:** 1=True 0=False 2=Not sure

*#Answers on previous fake news to be communicated to each respondent: StopCovid does not detect people, but Bluetooth devices. StopCovid does not track movements (no geo-location, no access to GPS data). StopCovid does not trace contacts between people. At no time is the name of the owners or users of this device requested or exchanged. The only information that goes from the phone to the central authority is the proximity for at least 15 minutes and within one meter of two unique identifiers (such as "d911b9b2ce5a6aef"). The identifiers to be used are regularly provided by the central authority to smartphones, with a life span limited to a few minutes#*

**SCENARIO 2 “No, I have never heard about the StopCovid mobile app**

**15) What do you think the app is about?**

1. An app that gives advice and information on Covid-19
2. An app that helps to stop the spread of the virus
3. Do not know
4. Other

If "Other", specify: ________________________

*#The interviewer quickly describes the app: It is a mobile app that allows warning people who have been close to a person who has tested positive, so that they can be taken care of as soon as possible.The app checks via Bluetooth whether the smartphone has been in close proximity (less than 1 meter) for more than 15 minutes with another smartphone of a person tested positive who owns the app. The app suggests what to do if contact is established through a notification. It works transparently and anonymously: no personal data is collected. There is just the exchange of a pseudo-identifier renewed every 15 minutes which does not allow the person to be identified. It is supported by the French government and was developed by a group of public and private actors coordinated by the French research institution Inria#*

**16) Now that you know the app, would you download it?** 1=YES 0=N0 2=NOT SURE/DO NOT KNOW

| **IF “YES, I WOULD DOWNLOAD THE APP”** | **IF “NO, I WOULD NOT DOWNLOAD THE APP” OR IF “I AM NOT SURE, I DO NOT KNOW”** |
|---|---|
| **16.1) Why?**<br>1. Out of curiosity<br>2. To protect my family, others and myself from possible infection<br>3. It was the government that advised the downloading of the app<br>4. The app could be useful to contain the spread of the virus in general<br>5. Am afraid of the virus and all strategies are good to avoid it<br>6. Am reassured the app is anonymous<br>7. Other<br>If "Other", specify: ____________________ | **16.2) Why? Why aren't you sure you want to download it?**<br>1. Cannot see the interest or need<br>2. Do not like the general idea of this app<br>3. Do not know how it works, did not get enough information on the app<br>4. Am suspicious of this type of app<br>5. Do not trust, because do not know who is offering this app: industry etc.<br>6. Not sure about the security of the data, fear of being geo-located<br>7. My family and friends have discouraged me from downloading it<br>8. No space on my phone or it is not powerful enough to have an extra app (battery, Bluetooth)<br>9. Do not carry my phone with me at all times |

| | |
|---|---|
| | 10. Do not use public transportation and/or do not go out much in public places (do not come into contact with strangers)<br>11. It does not seem to be effective (too few people use it)<br>12. Heard negative feedback on this app<br>13. Do not really have time to think about it<br>14. By negligence, not concerned<br>15. Other<br>If "Other", specify:____________________<br><br>**16.3) What could make you change your mind and download the app?**<br>1. Have a better phone<br>2. Learn more about the app (with better communication, for example)<br>3. Have positive feedback on its usefulness<br>4. Having cases of COVID-19 among my family and friends<br>5. Be even more reassured about its transparency and the protection of anonymity (clearer information on the transmission of information)<br>6. Many more people who use it<br>7. Know that the app is used by a majority of the people around me (e.g. other students that I may come across)<br>8. Nothing would change my mind<br>9. Other<br>If "Other, specify:_____________________ |

For both groups of SCENARIO 2

**12) In your opinion, what could help diffuse the app more widely?**

1. A better communication strategy (more visibility, clearer messages)
2. Providing better technical features (battery, Bluetooth, space on the phone)
3. An explosion of Covid-19 cases
4. Making it mandatory
5. Do not know
6. Other

If "Other", specify: ________________________

**13) Would you advise those around you to download the app?** 1=YES 0=NO 2=DO NOT KNOW

**14) Which of the following statements are true or false? You can also answer that you are not sure of the answer.**

**14.1) StopCovid geo-locates you and tracks your movements:** 1=True 0=False 2=Not sure

**14.2) StopCovid collects your contacts and knows their names (telephone directory, contacts on social networks...):** 1=True 0=False 2=Not sure

**14.3) StopCovid detects people around you and knows their names (physical contacts):** 1=True 0=False 2=Not sure

**14.4) StopCovid has access to your personal data and communicates them:** 1=True 0=False 2=Not sure

*#Answers on previous fake news to be communicated to each respondent: StopCovid does not detect people, but Bluetooth devices. StopCovid does not track movements (no geo-location, no access to GPS data). StopCovid does not trace contacts between people. At no time is the name of the owners or users of this device requested or exchanged. The only information that goes from the phone to the central authority is the proximity for at least 15 minutes and within one meter of two unique identifiers (such as "d911b9b2ce5a6aef"). The identifiers to be used are regularly provided by the central authority to smartphones, with a life span limited to a few minutes.#*
